# Clinical characteristics associated with COVID-19 severity in California

**DOI:** 10.1101/2020.03.27.20043661

**Authors:** Samuel J. S. Rubin, Samuel R. Falkson, Nicholas Degner, Catherine Blish

**Affiliations:** Department of Medicine, Stanford University School of Medicine, Stanford, CA, USA; Chan-Zuckerberg Biohub, San Francisco, CA, USA

## Abstract

Given the rapidly progressing COVID-19 pandemic, this report on a US cohort of 54 COVID-19 patients from Stanford Hospital and data regarding risk factors for severe disease obtained at initial clinical presentation is of high importance and is immediately clinically relevant. We identified low presenting oxygen saturation as predictive of severe disease outcomes, such as diagnosis of pneumonia, acute respiratory distress syndrome (ARDS), and admission to the ICU, and also replicated data from China suggesting a link between hypertension and disease severity. Clinicians will benefit by tools to rapidly risk stratify patients at presentation by likelihood of progression to severe disease.

## Introduction

The coronavirus disease 2019 (COVID-19) pandemic caused by severe acute respiratory syndrome coronavirus 2 (SARS-CoV-2) poses a global threat. Disease severity varies widely from mild (81%), requiring hospitalization (12%), to death (1.8-3.4%) (1).

The growing number of US COVID-19 cases is expected to tax the capacity of health care delivery systems. Analysis of clinical characteristics at time of patient presentation associated with disease severity is immediately useful. Recognizing the rapid utility of such data, we evaluated clinical characteristics and disease course of patients with confirmed COVID-19 at Stanford Hospital.

## Methods

With approval of the Stanford Institutional Review Board, patient charts were analyzed if they were diagnosed with COVID-19 by RT-PCR, received care at Stanford Hospital by March 16, 2020, and had past medical history documentation. Statistical analyses were conducted in Microsoft Excel and R.

## Results

Of 54 patients analyzed, the median age was 53.5 years (IQR, 32.75; range, 20–91), 53.7% (29) were male, 18 were inpatients, and 36 were outpatients (Table 1). Based on chart documentation of past medical history, 14 had hypertension, 13 had hyperlipidemia, and 7 had diabetes (6 type 2, 1 type 1).

**Table 1.**
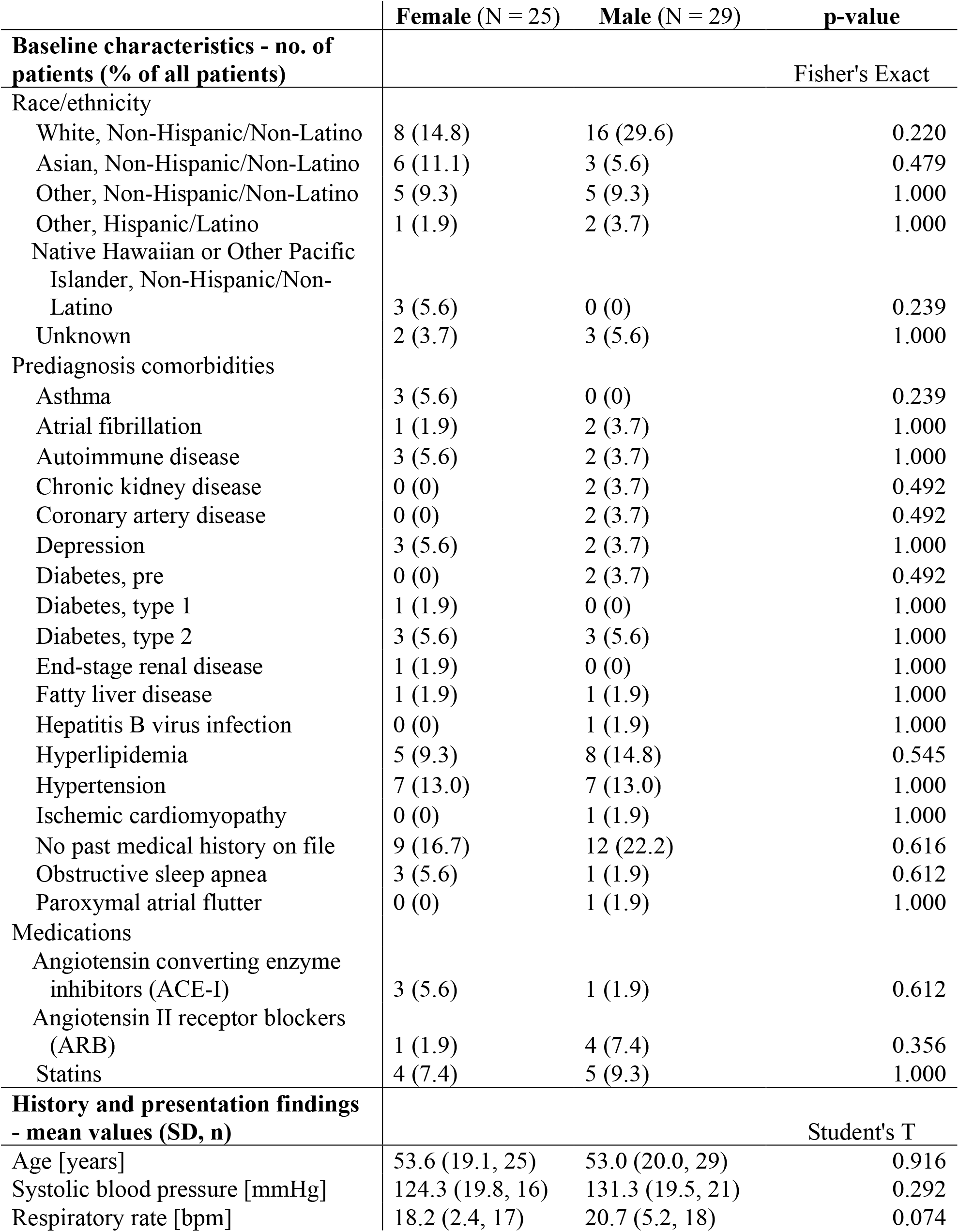
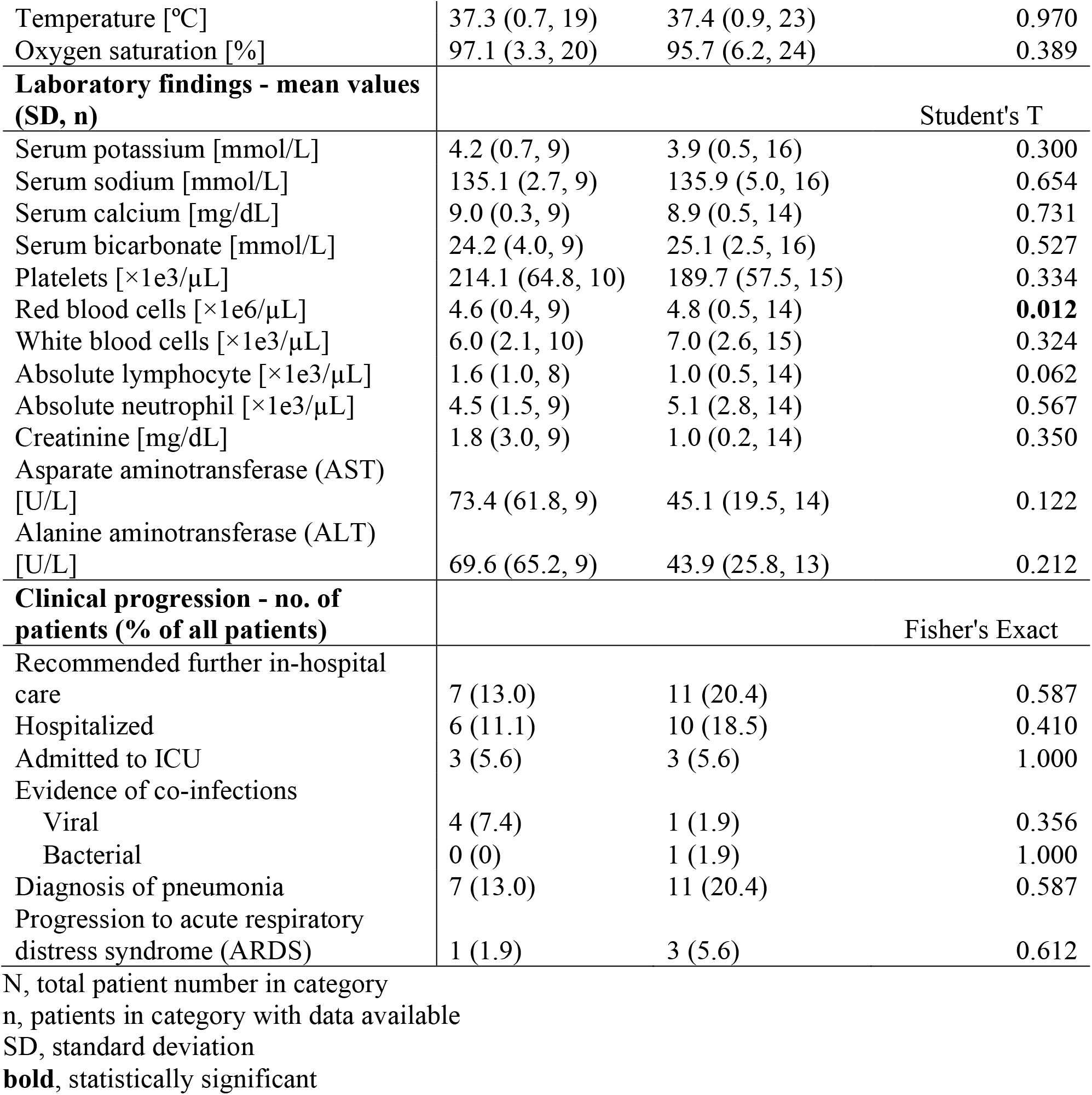
Clinical characteristics of 54 patients with COVID-19 in California.

Consistent with previous COVID-19 reports, the most common prediagnosis comorbidity in 24 patients (25.9%) was hypertension; this rate does not significantly differ from the US prevalence (Fisher’s exact test, p = 0.671) (2, 3). Angiotensin-converting enzyme inhibitor (ACE-I) or angiotensin II receptor blocker (ARB) use in 9 patients (16.7%) was also not significantly different from the US prevalence (Fisher’s exact test, p = 1.000) (4).

In univariate analysis, age, lower oxygen saturation at initial examination, and hypertension were significantly associated with recommendation for further hospital care, lower oxygen saturation was associated with admission to ICU, age and lower oxygen saturation were associated with diagnosis of pneumonia, and lower oxygen saturation and hypertension were associcated with progression to acute respiratory distress syndrome (ARDS) (Table 2). Use of ACE-I or ARB was not significantly associated with recommendation for further hospital care, admission to ICU, diagnosis of pneumonia, or progression to ARDS. When analyzed by logistic regression to control for age, the only factor independently significantly associated with recommendation for further in-hosptial care, diagnosis of pneumonia, and progression to ARDS was initial oxygen saturation measurement as a continuous variable.

**Table 2.** Correlates of clinical progression.

|  | **Univariate**<br>**p-value** | **Multivariate**<br>**p-value** |
| --- | --- | --- |
| **Recommended further in-hospital care** |  |  |
| Age* | **0.003** | 0.926 |
| Oxygen saturation at history and presentation* | **4.730E-04** | **0.010** |
| History of hypertension<sup>†</sup> | **0.024** | 0.138 |
| Use of angiotensin converting enzyme inhibitors (ACE-I)<sup>†</sup> | 1.000 | 0.339 |
| Use of angiotensin II receptor blockers (ARB)<sup>†</sup> | 0.057 | 0.239 |
| **Admission to ICU** |  |  |
| Age* | 0.204 | 0.879 |
| Oxygen saturation at history and presentation* | **6.667E-04** | 0.067 |
| History of hypertension<sup>†</sup> | 0.173 | 0.268 |
| Use of angiotensin converting enzyme inhibitors (ACE-I)<sup>†</sup> | 1.000 | 0.995 |
| Use of angiotensin II receptor blockers (ARB)<sup>†</sup> | 0.459 | 0.433 |
| **Diagnosis of pneumonia** |  |  |
| Age* | **0.027** | 0.862 |
| Oxygen saturation at history and presentation* | **0.001** | **0.026** |
| History of hypertension<sup>†</sup> | 0.512 | 0.186 |
| Use of angiotensin converting enzyme inhibitors (ACE-I)<sup>†</sup> | 0.289 | 0.994 |
| Use of angiotensin II receptor blockers (ARB)<sup>†</sup> | 1.000 | 0.075 |
| **Progression to acute respiratory distress syndrome** |  |  |
| Age* | 0.151 | 0.578 |
| Oxygen saturation at history and presentation* | **3.476E-05** | **0.029** |
| History of hypertension<sup>†</sup> | **0.049** | 0.094 |
| Use of angiotensin converting enzyme inhibitors (ACE-I)<sup>†</sup> | 1.000 | 0.997 |
| Use of angiotensin II receptor blockers (ARB)<sup>†</sup> | 0.330 | 0.467 |

\* Two-tailed homoscedastic Student's T test for univariate analysis
<sup>†</sup> Fisher's exact test for univariate analysis
**bold**, statistically significant

## Discussion

Clinical characteristics of US COVID-19 patients and factors from initial presentation that associate with disease severity were identified. Lower oxygen saturation at presentation was independently significantly associated with measures of disease severity, and thus may serve as a useful indicator of potential disease progression. Additionally, history of hypertension predisposed patients to worse outcomes such as ARDS, although not independently of age, consistent with previous reports (2).

Several factors, including age-related changes in the immune system and other physiological processes could contribute to COVID-19 disease severity. While hypertension is associated with COVID-19 morbidity, it is not independent of age; thus further study is needed to elucidate the extent to which hypertension and/or dysregulation of the renin-angiotensin-aldosterone system (RAAS) contributes to COVID-19 pathogenesis. The idea that RAAS dysregulation may influence disease progression warrants further exploration into the usefulness of ACE-I and/or ARB therapies in COVID-19 management.

The potential role of the RAAS system in COVID-19 pathogenesis stems from mounting sequence and structural evidence indicating entry of SARS-CoV-2 via interaction of its spike protein with its human host receptor angiotensin-converting enzyme 2 (ACE2) (5). It is difficult to predict the effects of RAAS blockade in treatment of COVID-19, as it may increase expression of ACE2, with known anti-inflammatory and pulmonary protective properties, yet simultaneously promote viral entry (6). However, RAAS blockade may also increase soluble ACE2, which could serve as a decoy receptor protecting against viral entry.

In our study, history of ACE-I or ARB use did not affect diagnosis rate or predispose patients to worse disease outcomes. However, our study is underpowered to draw definitive conclusions from such negative data. We hope these initial findings motivate larger studies intended to characterize RAAS blockade in the COVID-19 setting.

Limitations of this study include lack of some data on disease progression due to retrospective design and potential selection bias of patients with severe disease more likely to have SARS-CoV-2 testing and extensive charting. However, this US data is immediately clinically relevant given the rapidly evolving pandemic and will help clinicians identify and treat patients most at risk of severe disease.

## Data Availability

De-identified data is available from the corresponding author upon reasonable request.

## Acknowledgements

We thank Arjun Rustagi, MD, PhD, Vivek Bhalla, MD, and Glenn Chertow, MD, MPH of Stanford University School of Medicine, USA, and Andrew Michael South, MD, MS of Wake Forest School of Medicine, USA for helpful discussions. No compensation was received for their roles in the study.

## Notes

**Conflicts of Interest:** None to disclose.

### Competing Interest Statement

The authors have declared no competing interest.

### Funding Statement

No funding received.

## References

1. Bialek S, Boundy E, Bowen V, Chow N, Cohn A, Dowling N, Ellington S, Gierke R, Hall A, MacNeil J, Patel P, Peacock G, Pilishvili T, Razzaghi H, Reed N, Ritchey M, Sauber-Schatz E. Severe Outcomes Among Patients with Coronavirus Disease 2019 (COVID-19) — United States, February 12–March 16, 2020. MMWR Morb Mortal Wkly Rep 2020. doi: 10.15585/mmwr.mm6912e2

2. Wu C, Chen X, Cai Y, Xia J, Zhou X, Xy S, Huang H, Zhang L, Zhou X, Du C, Zhang Y, Song J, Wang S, Chao Y, Yang Z, Xu J, Zhou X, Chen D, Xiong W, Xu L, Zhou F, Jiang J, Bai C, Zheng J, Song Y. Risk Factors Associated With Acute Respiratory Distress Syndrome and Death in Patients With Coronavirus Disease 2019 Pneumonia in Wuhan, China. JAMA Intern Med 2020. doi:10.1001/jamainternmed.2020.0994

3. Hypertension Prevalence and Control Among Adults: United States, 2016–2016. Hyattsville, MD: National Center for Health Statistics; 2017. NCHS data brief, no 289.

4. Benjamin EJ, Virani SS, Callaway CW, Chamberlain AM, Chang AR, Cheng S, Chiuve SE, Cushman M, Delling FN, Deo R, de Ferranti SD, Ferguson JF, Fornage M, Gillespie C, Isasi CR, Jimenez MC, Jordan LC, Judd SE, Lackland D, Lichtman JH, Lisabeth L, Liu S, Longenecker CT, Lutsey PL, Mackey JS, Matchar DB, Matsushita K, Mussolino ME, Nasir K, O’Flaherty M, Palaniappan LP, Pandey A, Pandey A, Pandey DK, Reeves MJ, Ritchey MD, Rodriguez CJ, Roth GA, Rosamond WD, Sampson UKA, Satou GM, Shah SH, Spartano NL, Tirschwell DL, Tsao CW, Voeks JH, Willey JZ, Wilkins JT, Wu JHY, Alger HM, Wong SS, Muntner P. Heart Disease and Stroke Statistics—2018 Update: A Report From the American Heart Association. Circulation 2018. doi:10.1161/CIR.0000000000000558

5. Wan Y, Shang J, Graham R, Baric RS, Li F. Receptor recognition by novel coronavirus from Wuhan: An analysis based on decade-long structural studies of SARS. J Virol. 2020.

6. Ferrario CM, Jessup J, Chappell MC, Averill DB, Brosnihan KB, Tallant EA, Diz DI, Gallagher PE. Effect of Angiotensin-Converting Enzyme Inhibition and Angiotensin II Receptor Blockers on Cardiac Angiotensin-Converting Enzyme 2. Circulation 2005. doi:10.1161/CIRCULATIONAHA.104.510461

